# The Role of Physical Environmental Characteristics and Intellectual Disability in Conduct Problem Trajectories Across Childhood: A Population-Based Cohort Study

**DOI:** 10.1101/2021.09.13.21263494

**Authors:** Alister Baird, Efstathios Papachristou, Angela Hassiotis, Eirini Flouri

## Abstract

**Background:** The paucity of research investigating the role of the physical environment in the developmental progression of conduct problems and the potential moderating effects of intellectual disability (ID) is surprising, given the clinical relevance of elucidating environmental determinants of disruptive behaviours.

**Aims:** To use data from a large UK cohort study to assess associations between physical environmental exposures, ID, and conduct problem trajectories.

**Method:** The sample included 8,168 Millennium Cohort Study children (1.9% with ID). Multilevel growth curve modelling was used to examine the role of physical environment characteristics in the developmental trajectories of conduct problems after adjustments for ID status.

**Results:** Exposure to external environmental domains was not associated with differences in children’s conduct problems across development. Alternatively, internal aspects of the household environment: spatial density (b = 0.40, *p*<.001) and damp problems (b = 0.14, *p*<.001) were both significantly associated with increased trajectories. Various individual and familial covariates were positively associated with conduct problems over time, including: presence of ID (b = 0.96, *p*<.001), autism spectrum disorder (b = 1.18, *p*<.001), male sex (b = 0.26, *p*<.001), poverty (b = 0.19, *p*<.001), maternal depression (b = 0.65, *p*<.001), and non-nuclear family structure (b = 0.35, *p*<.001). Positive ID status appeared to moderate the effects of internal household spatial density, reporting a non-linear negative association with spatial density and conduct problems across development (b = -1.08, *p*<.01).

**Conclusions:** Our findings highlight the potential harmful consequences of poor internal residential conditions on children’s development of disruptive behaviours.

## Introduction

Interest in the role of children’s early physical environment on neurodevelopmental and socio-cognitive outcomes has increased in recent decades, evidenced by reviews examining its effects on well-being^1,2,3^, mental health^4,5^, and development^6^. Physical domains explored are diverse, ranging from nature exposure and meteorology, to architectural design. Previous studies have highlighted the valence of physical environmental aspects such as exposure to natural greenspaces^7^ and air particulate matter^8^ on childhood psychological and neurophysiological outcomes. Considering that challenging behaviours have been associated with a wide range of negative long-term consequences, ranging from high societal economic costs^9^, to reduced life satisfaction and disintegration of social connections^10^, it is surprising that there is an aperture of current literature examining the role of the physical environment in the developmental progression of childhood conduct problems. Moreover, it is yet to be examined whether the influence of the physical environment on disruptive behavior symptom trajectories is more severe for children with intellectual disability (ID) than typically developing children. The global prevalence of ID has been estimated at 1.37%^11^, with the frequency of comorbid challenging and aggressive behaviours in children ranging between 48% - 94%^12^. Physical environmental characteristics such as air pollution and urbanicity^13,14^ have been previously positively associated with conduct problems, whilst exposure to greenspaces has been negatively associated with these problems in both typically developing children^15,16^, and children with autism spectrum disorder^17^ (ASD). Although previous epidemiological work has explored the varied nature of conduct problems in children with cognitive difficulties^18^, very few systematic reviews examine environmental effects on the trajectories of disruptive behaviours in children with ID and neurodevelopmental disorders (NDDs) more broadly. Considering that the impact of physical environmental domains may be more salient in early neurodevelopmental stages^19^, and that consequences of childhood aggression are severe and persistent across the lifespan^20-26^, this study has the potential to illuminate the role of external and interior physical environmental features in the developmental trajectories of children’s conduct problems.

## Method

### Sample

The sample was drawn from the Millennium Cohort Study (MCS: https://cls.ucl.ac.uk/cls-studies/millennium-cohort-study/^27^), a UK population-based longitudinal birth cohort study. We used data collected from participants at MCS waves 2, 3, 4 and 5, with children aged, 3, 5, 7, and 11 years, respectively. In total 19,243 families have participated in the MCS to date. The MCS employed a stratified sampling protocol which disproportionally recruited disadvantaged or ethnic minority families (see Plewis^28^). The analytic sample of this study (n = 8,168; 49% male) included children (singletons and first-born twins or triplets) with valid data on cognitive ability and the behavioral outcome measure (conduct problems) across assessments.

### Intellectual disability ascertainment

To identify children with intellectual disability, first we ran principal components analysis (PCA) on standardised cognitive ability assessments available at each wave to derive summary cognitive scores. Such synthesis of multiple well-validated assessments has been shown to produce an adequate metric capturing general cognitive ability^29^. Next, children scoring more than two standard deviations below the mean score for cognitive ability at two or more waves were classified with a positive ID status. Parental confirmation of special educational needs (SEN) statement was also used to provide a positive ID classification in situ of missing cognitive scores, children who received a SEN statement due to being classified as ‘gifted’, or ‘high IQ’, were excluded.

At MCS wave 2 (child age 3), the Bracken School Readiness Assessment-Revised^30^ (conceptual knowledge) (BSRA-R) and the BAS-II Naming Vocabulary test^31^ (expressive language ability) were used to assess cognitive ability. For MCS wave 3 (child age 5) cognition was assessed using the: BAS Naming Vocabulary, BAS Pattern Construction (spatial problem-solving ability) and BAS Picture Similarities (non-verbal reasoning). For MCS wave 4 (children aged 7) the age standardised pattern construction and word reading scores from the BAS-II were used as well as a shortened version of the Progress in Maths tests^32^ (National Foundation for education Research). At wave 5 (child age 11) cognitive ability was assessed with the BAS Verbal Similarities (verbal knowledge and reasoning capability) and the error scores on the spatial working memory (SWM) task (representative of executive functioning^33^) from the Cambridge Neuropsychological Test Automated Battery^34^ (CANTAB). In the analytic sample the prevalence rate for ID was 1.9% (N = 155).

### Conduct problems

*Conduct problems* at ages 3, 5, 7, and 11 years were measured using the parent-reported Strengths and Difficulties Questionnaire^35^ (SDQ). In MCS, the conduct problems subscale of the SDQ has been shown to have satisfactory internal reliability with Cronbach’s alphas ranging from 0.77 and 0.82 for ages 3 – 7 years^36^, and 0.68 for children aged 11^37^. In the analytic sample, Cronbach’s alpha values ranged from 0.53 to 0.60 across assessments. Importantly, the SDQ has been shown to be comparably valid across populations with and without ID^38,39^.

### External physical environmental exposures

*Greenspace* was assessed in the MCS using ward-level greenspace data from the UK, estimated with data from the Generalized Land Use Database^40^ (GLUD) and from the Coordination of information on the Environment^41^ (CORINE; EEA). Regression models predicted GLUD percentage greenspace per English ward which was adapted for use on the whole of the UK^42^ resulting in a linear greenspace metric reporting the deciles of percentage of greenspace within wards, with higher scores reflective of more greenspace. Information on children’s *access to private garden space* was also available. Data on *pathogenic air particulate matter* in the UK was obtained from the Multiple Environmental Deprivation Index (MEDIx) and linked to children’s residential addresses. Annual population weighted mean concentrations of nitrogen dioxide (NO_2_) were taken between 1999 – 2003 from 2001 UK Census Areas Statistics (CAS) ward with annual means converted into deciles (higher scores representative of increased NO_2_) before being linked to MCS waves^43^. Data from the National Office of Statistics (ONS) was used to assess *urbanicity or rurality* of children’s residential geography^44^ at each included wave.

### Interior physical environmental exposures

*Home spatial density* was measured by extracting parent reported data on the total number of people residing in each household at each wave, divided by the total number of rooms. Parents reported information on *residential damp problems* per wave and this was transformed into a binary variable indicating presence or not of household damp.

### Covariates

Adjustment for a variety of time-invariant and time-varying covariates were made in our analyses. Time-invariant covariates included *ethnicity* (white or non-white), *sex*, and *autism spectrum disorder (ASD) status*. ASD confirmation was obtained from parental and teacher questionnaires specifying whether the child has a previous ASD diagnosis across at least 2 assessment waves. The time-varying covariates measured at each assessment wave included, *age (in years), poverty* (household income below 60% median national income obtained from the Organisation for Economic Co-operation and Development: OECD), *maternal depression*, and *family structure*. Maternal depression was measured using scores from the Kessler K6 depression scale^45^, a 6-item Likert response questionnaire, measuring psychological distress related to symptoms of depression and anxiety. We used previously validated diagnostic cut-off values greater than 13 as representative of serious mental illness (SMI) and depression^45^. Nuclear family structure was defined as the presence of both biological parents in the household.

### Statistical analysis

We initially examined whether children in the analytic sample differed from the remaining MCS sample in terms of their sociodemographic characteristics, exposure to external and interior physical environmental exposures, and conduct problem levels. Next, growth curve models (GCM) were fitted as random intercept multilevel models (MLM) to investigate the impact of external (greenspace, access to private garden space, air pollution, and urbanicity) and interior (home spatial density and residential damp problems) environmental measures on the developmental trajectories of conduct problems. The MLM had three levels; repeatedly measured data points for individuals (Level1) were treated as being nested within children (Level 2), which were in turn, were nested within electoral wards (Level 3). Children’s age was measured in years and was centred around the grand mean (6.71 years). Parameterised in this way, the effect of the predictor variables reflect mean differences at approximately age 7 years. We included an additional quadratic term for age in all models to explore the temporal linearity of conduct problems. Missing data in covariates were multiply imputed (MI) using chained equations (MICE) under the assumption that data is missing at random (MAR). Percentages of missing data ranged between 0.02 – 3.38%. A total of 20 datasets were imputed^46^. The main part of the analyses included two MLM: The first model (Model A) sought to examine the impact of external and interior physical environmental exposures on the trajectories of conduct problems after adjustments for ID status. The second model (Model B) examined the effects of the exposure variables on the outcome after adjustments for the time-varying (maternal psychological distress, socioeconomic disadvantage, family structure) and time-invariant cofounders (sex, ethnicity, ASD status). For both models we calculated the associated intra-class correlation coefficients (ICC) to estimate the variance in the trajectories that is attributable to the clustering of the variables within higher levels (Levels 2 and 3). Additional models including all two-way and three-way interaction terms between ID status, physical environmental measures, and age, were run to assess if the environmental influences on trajectories conduct problems varied dependent on children’s ID classification. We applied a stringent p-value significance criterion (^*^ = *p*<0.01) to all models to adjust for multiple comparisons.

Because the MCS sample is disproportionately stratified due to the intentional over-sampling of sub-groups of the population all models were accordingly adjusted by including appropriate stratification variables. Each country had two strata: advantaged and disadvantaged. England had an additional one for areas with high percentage of ethnic minorities. We also adjusted for the clustered MCS sample at electoral ward level but also for attrition and non-response rates by incorporating study-specific weights into our models. All analyses were run in Stata SE 16.1^47^.

## Results

### Bias analysis

First, we explored differences between children in the analytic (N = 8,168) and non-analytic sample (N = 11,075, Table 1). Children in the analytic sample lived in greener, less air polluted neighbourhoods and were more likely to have access to a private garden and to reside in more rural environments. The profile of their indoor home environments also differed, with residential homes being less spatially dense and with fewer damp problems. Significantly higher proportions of the non-analytic sample had income below the 60% medium national level, psychologically distressed mothers, ASD, and lived in non-nuclear family structures.

**Table 1.** Bias analysis between analytic and non-analytic samples.

|  | Analytic sample (n = 8168) |  | Non-analytic sample (n = 11,075) |  | Test |
| --- | --- | --- | --- | --- | --- |
| Continuous variables | n | M (SD) | n | M (SD) | F |
| SDQ conduct scores Sweep 2 | 8168 | 2.70 (2.01) | 6602 | 2.99 (2.13) | 75.05** |
| SDQ conduct scores Sweep 3 | 8168 | 1.40 (1.42) | 6577 | 1.67 (1.61) | 121.82** |
| SDQ conduct scores Sweep 4 | 8168 | 1.27 (1.47) | 5304 | 1.59 (1.65) | 132.99** |
| SDQ conduct scores Sweep 5 | 8168 | 1.29 (1.50) | 4630 | 1.56 (1.69) | 87.22** |
| Greenspace Sweep 2 | 8167 | 4.49 (2.67) | 7422 | 4.07 (2.67) | 206.44** |
| Greenspace Sweep 3 | 8167 | 4.78 (2.69) | 7078 | 4.15 (2.70) | 206.46** |
| Greenspace Sweep 4 | 8166 | 4.84 (2.69) | 5689 | 4.17 (2.73) | 202.55** |
| Greenspace Sweep 5 | 8164 | 4.91 (2.69) | 5116 | 4.15 (2.72) | 249.94** |
| Air pollution (NO<sub>2</sub>) Sweep 2 | 8167 | 6.15 (2.91) | 7422 | 6.77 (3.06) | 166.79** |
| Air pollution (NO<sub>2</sub>) Sweep 3 | 8167 | 6.66 (3.08) | 7078 | 6.66 (3.08) | 132.34** |
| Air pollution (NO<sub>2</sub>) Sweep 4 | 8166 | 6.05 (2.91) | 5689 | 6.65 (3.09) | 132.66** |
| Air pollution (NO<sub>2</sub>) Sweep 5 | 8164 | 5.99 (2.90) | 5116 | 6.79 (3.06) | 227.77** |
| Household spatial density Sweep 2 | 8166 | 0.72 (0.24) | 7255 | 0.82 (0.32) | 448.91** |
| Household spatial density Sweep 3 | 8157 | 0.73 (0.24) | 6967 | 0.82 (0.31) | 449.96** |
| Household spatial density Sweep 4 | 8154 | 0.74 (0.26) | 5573 | 0.85 (0.32) | 467.08** |
| Household spatial density Sweep 5 | 8120 | 0.88 (0.28) | 4814 | 1.02 (0.35) | 594.22** |
| Child Age (years) Sweep 2 | 8168 | 3.17 (0.19) | 7422 | 3.20 (0.30) | 55.81** |
| Child Age (years) Sweep 3 | 8168 | 5.28 (0.24) | 7077 | 5.30 (0.26) | 24.68** |
| Child Age (years) Sweep 4 | 8168 | 7.22 (0.24) | 5676 | 7.26 (0.27) | 87.82** |
| Child Age (years) Sweep 5 | 8168 | 11.16 (0.32) | 5119 | 11.19 (0.34) | 27.37** |
| Categorical variables | n | % | n | % | Chi<sup>2</sup> |
| Access to private garden Sweep 2 | 8107 | 94.99 | 7198 | 89.25 | 176.85** |
| Access to private garden Sweep 3 | 8096 | 95.81 | 6051 | 90.38 | 167.82** |
| Access to private garden Sweep 4 | 8095 | 95.96 | 4497 | 90.95 | 132.24** |
| Access to private garden Sweep 5 | 8083 | 96.67 | 3736 | 91.70 | 135.45** |
| Urban residence Sweep 2 | 8167 | 77.01 | 7422 | 81.92 | 57.28** |
| Urban residence Sweep 3 | 8167 | 75.84 | 7078 | 81.63 | 75.43** |
| Urban residence Sweep 4 | 8166 | 75.25 | 5689 | 81.28 | 70.39** |
| Urban residence Sweep 5 | 8164 | 74.31 | 5116 | 81.43 | 90.10** |
| Household damp problems Sweep 2 | 8168 | 12.82 | 7279 | 16.06 | 32.91** |
| Household damp problems Sweep 3 | 8165 | 11.87 | 6989 | 14.48 | 22.61** |
| Household damp problems Sweep 4 | 8166 | 13.38 | 5596 | 16.24 | 21.83** |
| Household damp problems Sweep 5 | 8142 | 16.20 | 4872 | 18.45 | 10.94** |
| Below poverty line Sweep 2 | 8151 | 25.09 | 7248 | 41.96 | 493.47** |
| Below poverty line Sweep 3 | 8164 | 25.12 | 6971 | 44.33 | 618.27** |
| Below poverty line Sweep 4 | 8165 | 22.79 | 5672 | 40.57 | 502.82** |
| Below poverty line Sweep 5 | 8168 | 16.12 | 5119 | 38.97 | 877.89** |
| Maternal psychological distress Sweep 2 | 7670 | 2.71 | 5920 | 4.53 | 32.57** |
| Maternal psychological distress Sweep 3 | 8070 | 2.86 | 6262 | 4.50 | 27.51** |
| Maternal psychological distress Sweep 4 | 8097 | 3.11 | 5066 | 4.46 | 16.20** |
| Maternal psychological distress Sweep 5 | 7732 | 4.45 | 3973 | 8.38 | 74.48** |
| Atypical family structure Sweep 2 | 8153 | 15.80 | 7323 | 25.00 | 203.16** |
| Atypical family structure Sweep 3 | 8164 | 20.22 | 7071 | 30.39 | 209.29** |
| Atypical family structure Sweep 4 | 8164 | 23.80 | 5683 | 33.71 | 163.86** |
| Atypical family structure Sweep 5 | 8164 | 30.56 | 5115 | 39.47 | 111.29** |
| Male | 8168 | 48.98 | 11075 | 53.18 | 33.18** |
| Autism Spectrum Disorder | 7958 | 0.44 | 7539 | 1.25 | 30.55** |
| Ethnicity White | 8168 | 90.45 | 11050 | 75.61 | 698.22** |
| England-Advantaged | 2698 | 33.03 | 2130 | 19.23 | 476.25** |
| England-Disadvantaged | 1971 | 24.13 | 2834 | 25.59 | 5.34* |
| England-Ethnic Minority | 557 | 6.82 | 2034 | 18.37 | 537.89** |
| Wales-Advantaged | 381 | 4.66 | 451 | 4.07 | 3.99* |
| Wales-Disadvantaged | 792 | 9.70 | 1136 | 10.26 | 1.64 |
| Scotland-Advantaged | 562 | 6.44 | 619 | 5.59 | 6.08* |
| Scotland-Disadvantaged | 407 | 4.98 | 784 | 7.08 | 35.57** |
| Northern Ireland-Advantaged | 350 | 4.29 | 373 | 3.37 | 10.93** |
| Northern Ireland-Disadvantaged | 486 | 5.95 | 714 | 6.45 | 1.99 |

Note. \*p < .05, \*\*p < .01

### Physical environment and intellectual disability

Next, MLMs were run to examine the impact of physical environmental exposures and ID on trajectories of conduct problems across childhood. Model A included physical environmental exposures and ID status as the only explanatory variables. The ICC estimates suggested that individual child differences (Level 2) explained 51.5% of the variance in conduct problem trajectories, while commonalities of children residing within the same electoral wards (level 3) explained a mere 1.7% of the variation in the outcome. The unstandardised regression estimates of the model are summarised in Table 2. The results showed that children with ID (b = 1.270, SE = 0.176, *p*<.001), and those residing in houses with higher residential spatial density (b = 0.356, SE = 0.062, *p*<.001) and with household damp (b = 0.163, SE = 0.033, *p*<.001) had more conduct problems at around 7 years (the centred intercept). In contrast, none of the exterior environmental characteristics considered were significantly associated with the conduct problem trajectories. The regression estimates of the linear (b = -0.217, SE = 0.005, *p*<.001) and quadratic (b = 0.052, SE = 0.001, *p*<.001) age terms indicated that, across childhood, conduct problems decreased in a nonlinear fashion.

**Table 2:** Model A (minimally adjusted GCM predicting SDQ conduct scores, *N* = 8168).

| Fixed effects | Coefficient (SE) | 95% CI |
| --- | --- | --- |
| Age | -0.217 (0.005)** | [-0.226, -0.208] |
| Age<sup>2</sup> | 0.052 (0.001)** | [0.049, 0.054] |
| Intellectual disability | 1.270 (0.176)** | [0.925, 1.614] |
| Greenspace (ward decile) | -0.003 (0.009) | [-0.021, 0.015] |
| Air pollution (NO<sub>2</sub>) | -0.021 (0.010) | [-0.040, -0.002] |
| Access to private garden | -0.236 (0.146) | [-0.523, 0.051] |
| Urban residence | 0.107 (0.057) | [-0.005, 0.219] |
| Home spatial density | 0.356 (0.062)** | [0.234, 0.477] |
| Damp / condensation | 0.163 (0.033)** | [0.098, 0.227] |
| Constant | 1.148 (0.191)** | [0.774, 1.523] |
| Random effects | Estimate (SE) | 95% CI |
| Level 3 (ward-level) |  |  |
| Intercept variance | 0.218 (0.025) | [0.174, 0.273] |
| Level 2 (Child level) |  |  |
| Slope (age) variance | 0.140 (0.005) | [0.131, 0.149] |
| Intercept variance | 1.120 (0.019) | [1.083, 1.159] |
| Intercept-slope variance | -0.329 (0.036) | [-0.398, -0.256] |

Note. Age was measured in years and grand mean centered (6.71 yrs).
For fixed effects: \* $p < .01$ , \*\* $p < .001$

After adjustments for the covariates (Model B), the ICC values associated with clustering within the second and third level were reduced to 48.2% and 1.3%, respectively, suggesting that a significant amount of variance in conduct problems trajectories was still accounted for by individual-child differences. Table 3 summarises the results of this MLM. Overall, after adjustments for ASD status, ethnicity, sex, family structure, poverty, and maternal depression, children with ID still had more conduct problem scores at around age 7 (b = 0.963, SE = 0.175, *p*<.000). Presence of damp (b = 0.143, SE = 0.031, *p*<.000) and high spatial density (b = 0.400, SE = 0.062, *p*<.000) were significantly associated with elevated levels of conduct problem after adjustments for confounding. As in Model A, we did not find evidence for an impact of greenspace, NO_2_ particulate matter, access to private gardens or urbanicity on conduct problem trajectories.

**Table 3:** Model B (fully adjusted GCM predicting SDQ conduct scores, *N* = 8168).

| Fixed effects | Coefficient (SE) | 95% CI |
| --- | --- | --- |
| Age | -0.224 (.005)** | [-0.234, -0.215] |
| Age<sup>2</sup> | 0.052 (.001)** | [0.049, 0.054] |
| Intellectual disability | 0.963 (.175)** | [0.620, 1.307] |
| Greenspace (ward decile) | -0.000 (0.009) | [-0.018, 0.017] |
| Air pollution (NO<sub>2</sub>) | -0.014 (0.009) | [-0.031, 0.004] |
| Access to private garden | -0.164 (0.130) | [-0.419, 0.091] |
| Urban residence | 0.071 (0.056) | [-0.039, 0.181] |
| Home spatial density | 0.400 (0.062)** | [0.277, 0.523] |
| Damp / condensation | 0.143 (0.031)** | [0.083, 0.203] |
| Maternal psychological distress | 0.650 (0.098)** | [0.458, 0.842] |
| Below poverty line | 0.190 (0.037)** | [0.117, 0.262] |
| Atypical family structure | 0.346 (0.036)** | [0.275, 0.416] |
| Male | 0.263 (0.035)** | [0.195, 0.331] |
| Autism Spectrum Disorder | 1.182 (0.311)** | [0.572, 1.793] |
| White ethnicity | 0.170 (0.068) | [0.036, 0.304] |
| Constant | 0.605 (0.181)* | [0.251, 0.959] |
| Random effects | Estimate (SE) | 95% CI |
| Level 3 (ward-level) |  |  |
| Intercept variance | 0.176 (0.022) | [0.138, 0.224] |
| Level 2 (Child level) |  |  |
| Slope (age) variance | 0.139 (0.005) | [0.131, 0.148] |
| Intercept variance | 1.058 (0.018) | [1.024, 1.094] |
| Intercept-slope variance | -0.356 (0.034) | [-0.420, -0.288] |

Note. Age was measured in years and grand mean centered (6.71 yrs).
For fixed effects: \* $p < .01$ , \*\* $p < .001$

Regarding the effect of covariates, those classified as ASD (b = 1.182, SE = 0.311, *p*<.000), males (b = 0.263, SE = 0.035, *p*<.000), those whose mothers were psychologically distressed (b = 0.650, SE = 0.098, *p*<.000) and those living in non-nuclear (b = 0.346, SE = 0.036, *p*<.000) or poorer families (b = 0.190, SE = 0.037, *p*<.000) had, on average, more conduct problems at around 7 years.

### Interactions between physical environment and intellectual disability

To assess if exposure to physical environmental aspects was differentially associated with conduct problem trajectories for children with or without ID, additional GCM models, each including all 2-way interaction terms and the 3-way interaction term between environmental exposures, ID status and age, were run (results summarised in Tables S1-S5 in the supplementary material). ID did not appear to modify associations between external physical environment exposures and conduct problems. However, the effect of the interaction term between ID and spatial density on conduct problems was statistically significant (b = -1.075, SE = 0.346, *p*<.01), suggesting that positive ID status and increased household spatial density was associated with decreases in conduct problems over time (Table 4).

**Table 4:** Spatial density x ID diagnosis growth curve interaction model predicting SDQ conduct scores (N = 8168).

| Fixed effects | Coefficient (SE) | 95% CI |
| --- | --- | --- |
| Age | -0.195 (0.010)** | [-0.215, -0.170] |
| Age<sup>2</sup> | 0.053 (0.001)** | [0.050, 0.055] |
| ID diagnosis | 1.882 (0.340)** | [1.118, 2.646] |
| Greenspace (ward decile) | -0.000, (0.009) | [-0.017, 0.017] |
| Air pollution (NO<sub>2</sub>) | -0.013 (0.009) | [-0.031, 0.004] |
| ID diagnosis x spatial density x Age | -0.055 (0.063) | [-0.178, 0.068] |
| Spatial density x Age | -0.040 (0.012)* | [-0.063, -0.017] |
| ID diagnosis x Age | 0.123 (0.061) | [0.006, 0.251] |
| ID diagnosis x Spatial density | -1.075 (0.346)* | [-1.753, -0.397] |
| Urban residence | 0.072 (0.056) | [-0.038, 0.182] |
| Access to private garden | -0.153 (0.130) | [-0.408, 0.102] |
| Home spatial density | 0.460 (0.063)** | [0.257, 0.495] |
| Damp / condensation | 0.145 (0.031)** | [0.085, 0.205] |
| Maternal psychological distress | 0.654 (0.098)** | [0.461, 0.846] |
| Below poverty line | 0.192 (0.037)** | [0.119, 0.264] |
| Atypical family structure | 0.348 (0.036)** | [0.278, 0.419] |
| Male | 0.263 (0.035)** | [0.195, 0.331] |
| Autism Spectrum Disorder | 1.156 (0.312)** | [0.543, 1.768] |
| White ethnicity | 0.165 (0.069)* | [0.031, 0.299] |
| Constant | 0.549 (0.182)* | [0.193, 0.905] |
| Random effects | Estimate (SE) | 95% CI |
| Level 3 (ward-level) |  |  |
| Intercept variance | 0.178 (0.022) | [0.139, 0.225] |
| Level 2 (Child level) |  |  |
| Slope (age) variance | 0.138 (0.005) | [0.129, 0.147] |
| Intercept variance | 1.058 (0.018) | [1.023, 1.093] |
| Intercept-slope variance | -0.351 (0.034) | [-0.417, -0.283] |

Note. Age was measured in years and grand mean centered (6.71 yrs).
For fixed effects: \* $p < .01$ , \*\* $p < .001$

### Sensitivity analysis

We investigated this association between spatial density, ID, and conduct problems further by conducting a sensitivity analysis to examine if the ethnicity of children with ID influenced conduct problem trajectories. We found that white ethnicity and increased spatial density interacted significantly to predict elevated SDQ conduct scores (b = 0.419, SE = 0.113, *p*<.000), however the 3-way interaction including ID did not (b = 0.906, SE = 0.670, *p* = 0.176), suggesting that whilst ethnicity (white / non-white) interacted with home crowdedness in predicting conduct problem development, ID diagnosis did not appear to moderate this relationship (Supplementary Table 6).

## Discussion

The role of physical environmental exposures on children’s socioemotional and neurodevelopment is currently not well understood, with previous calls for large scale multilevel interdisciplinary approaches to investigate its impacts being issued^48^. In this study, we used data from UK’s largest recent birth cohort to examine associations between various interior and external environmental aspects with conduct problems across childhood. Additionally, the role of ID as a potential moderator of these associations was examined to investigate if physical environment differentially effects children’s conduct problems dependent upon ID diagnosis, as previous commentary^49^ has suggested that the antecedent motivations and aetiology of disruptive behaviours in individuals with ID may not differ fundamentally from typically developing individuals.

In this sample, external physical environmental domains were not associated with conduct problems, whilst both home crowdedness and damp were both related to increased conduct problem scores across development. In-line with previous literature, multiple socio-economic and familial covariates were also associated with behavioural problems, including: maternal psychological distress^50^, poverty^51^, living within a non-nuclear family structure^52^, ethnicity^53^, and male sex^54^.

Our lack of significant associations between greenspace and conduct problems is incongruous with literature examining its effects on children’s psychobehavioural outcomes. For example, some of the available studies suggest advantageous effects of access to greenspace on children’s conduct problems^55-59^. Whilst our findings may be indicative of a true null association between greenspace exposure and child conduct problems, it may also be an artefact of the longitudinal nature of this sample, as previous studies have been primarily cross-sectional. It may also be attributable to inadequacies in the ward-level greenspace data available within the MCS, its reliance on geographical location and inability to assess the quality and frequency of children’s greenspace exposure has been highlighted previously^60^. These are critical components of greenspace contact^61^ and their omission may draw into question the validity of this metric. Whilst children’s access to private gardens did trend towards reductions in conduct problem trajectories in this study, it did not reach significance. Again, these results are discordant with previous studies which reported beneficial associations between access to private gardens and childhood conduct problems^62,63^. Due to safety concerns, children’s opportunity for autonomous play has diminished in recent decades^64-67^, potentially limiting their access to neighbourhood greenspaces and making private garden access a more salient measure of greenspace exposure.

Similarly, NO_2_ exposure was not correlated with conduct problems in our sample. This mirrors previous findings which found no correlation between NO_2_ particulate matter^68^ or elemental carbon attributed to traffic pollution (ECAT) on childhood conduct problems. Nonetheless, the available evidence on the associations between air pollution and behavioural outcomes is mixed, with a previous study reporting^23^ harmful associations between particulate matter less than 2.5 microns (PM_2.5_) and NO_2_ exposure, and increased risk of conduct disorders. This may suggest that cumulative exposure to air pollution grows during childhood, resulting in negative behavioural outcomes manifesting later in development, postliminary to the latest assessment used in our analysis (age 11). Recent research on the effects of ambient air pollution on children with ID - also using the MCS^69^ - reported that they are between 17% - 33% more likely to live in areas of high air pollution dependent on the particulate matter exposure measured. Neurotoxicity related to air pollutants has also been linked to the aetiology of NDDs^70,71^, and in a recent review of the current epidemiological evidence, Xu, Ha, and Basnet^72^ reported the harmful effects of exposure to various air pollutants (H_2_S, ozone, PM_10_, PM_2.5_, and NO_2_, among others) on neurodevelopment and psychobehavioural functioning in both adults and children. This is in accordance with recent research linking the presence of ultra-fine air particulate matter in the brain to neuroinflammatory and intrathecal inflammatory responses^73^, highlighting valid neurophysiological mechanisms for the aetiology of air pollution in childhood aggression.

We report no association between urbanicity or rurality of residential geography and conduct problems. Previous studies that have examined the relationship between urban/rural residence and aggression outcomes in children, reporting contradictory findings^74-78^. Urban environments have been shown to increase exposure to potentially harmful environmental influences (noise pollution, air pollution, overcrowding) that may place additional strain on cognitive and self-regulatory processes^79^, contributing to the aetiology of violent behaviour^80^. Urban inhabitants also have limited access to greenspaces^81^, which have been purported to operate as a protective buffer against harmful environmental exposures^82,83^, potentially compounding the negative effects of urban stimuli. It is worth considering if the possible therapeutic efficacy of greenspace exposure may be attributable to reduced exposure to harmful urban environmental stimuli^84^ rather than a direct benefit of nature experience itself.

Presence of damp in children’s homes was associated with increased SDQ conduct problems scores across development. Previous work using MCS data reported negative effects of children’s indoor environment on cognitive and behavioural processes such as self-regulation and conduct problems^85,86,60^. Increased damp can lead to toxic mould and poor air quality^87^ to which children are especially vulnerable^88^ and which may cause neuroinflammatory and/or neurotoxic responses^89,90^. Financial circumstances and poor interior physical conditions of residential homes are highly correlated^91^, it is therefore likely that homes with multiple structural deficits exacerbate there negative effects on children’s mental health. Future research should aim to elucidate the direct toxic neurophysiological effects of household damp and mould, and attempt to separate these from the influences of often comorbid low socioeconomic status that accompany occupation of deficient home environments.

Increased spatial density was associated with decreased conduct problem trajectories in children with a positive ID status. This finding is rather counterintuitive; previous research on spatial density and aggressive behaviour has primarily examined the influence of low-density vs high-density playground or classroom conditions in typically developing children, reporting disparate associations^92-95^. One contemporary study^96^ reported that residential overcrowding was correlated with increased teacher reported externalising behaviors. Parents in crowded households have been shown to be less attentive and patient^97^, potentially indicative of a habituation effect leading to inaccurate caregiver reported conduct problems. The paucity of research exploring the impact of crowding on onset and/or maintenance of conduct problems in children with ID makes it impossible to embed our findings within the wider literature. This association between spatial density and conduct problems in children with ID may be attributable to several causes: first, this analysis was underpowered due to the relatively small sample of children with ID in this cohort. Second, households with an elevated density of family members may convey additional benefits to children with ID, for example increased availability of support from proximal family members. Third, due to limited availability of complex spatial density metrics within the MCS, we adopted a one-dimensional measure of parental reported residential crowding, which may fail to capture the whole range of the spatial dynamics in the home. Finally, children with ID may spend a higher proportion of time in indoor environments due to the additional logistical complexity of undertaking activities outside of the home. This is especially relevant given contextual circumstances related to the COVID-19 pandemic pertaining to the additional risks and difficulties of complying to infection control measures^98^.

## Strengths and limitations

To our knowledge, this is the first study exploring the influences of children’s physical environment on conduct problem trajectories whilst examining moderating effects of ID diagnosis. A strength of this work is the large and diverse sample size facilitated by the MCS, even after exclusion of participants due to missing data. The longitudinal nature of this study also allows the examination of conduct problems across children’s early neurodevelopmental periods.

Several limitations are also present in the current study, and the results should be interpreted with these caveats in mind. One is the dependence on the accuracy of parental reported SDQ conduct scores, a metric likely to contain inherent biases^15^. Another is the stringency in how we derived our ID classification, which may have resulted in under-ascertainment, however due to inability to certify clinical ID diagnosis this was deemed appropriate, and the resulting ID prevalence in our sample is comparative to previous reported global prevalence rates^11^. It is also worth drawing attention to our bias analysis which evidenced consistent differences between our analytic and non-analytic sample across variables. The lack of diversity in physical environmental aspects that were included in the MCS data collection protocol may be considered a limitation of this work. Ambient road traffic noise for example, has been associated with elevated parent reported conduct problems in children^99,100^, therefore the inability to include potential additional confounding physical environmental influences in analysis may limit the external validity of findings.

## Conclusion

In conclusion, external physical environmental measures were not associated with childhood trajectories in this population; moreover, classification of ID did not appear to mediate these relationships. Residential crowdedness and damp problems were both associated with increased conduct problems during development. Investigation of the moderating influence of ID status reported significant interaction effects for home crowding only, reporting a negative non-linear association with children’s conduct problems trajectories. A dearth of research on the influence of spatial density on children with ID inhibits extrapolation of this finding, and caution in interpretation is warranted.

Additional individual, sociodemographic, and familial covariates such as: ID and ASD diagnosis, maternal psychological distress, poverty, living in a non-nuclear familial structure, and male sex were also significant predictors of increased conduct problems trajectories.

This work highlights the scarcity of contemporary research investigating the influence of physical environmental factors on the pathogenesis and progression of conduct problems in childhood NDDs. Previous work has outlined how NDD populations are disproportionately affected by health inequalities largely attributable to preventable environmental determinants^101^. Despite calls to improve mental ill-health of children over the previous decade^102^, systemic study into the harmful effects of children’s physical environment, and strategies to mitigate these preventable risks have not been conducted. Understanding the harmful and therapeutic influences of physical environmental domains can inform special educational policy and facilitate additional tools for clinicians and caregivers to alleviate a range of detrimental neurobehavioral outcomes, including conduct problems.

## Data Availability

The data that support the findings of this study are available from the corresponding author, [A.B.], upon reasonable request and approval from relevant research authorities.

## Data Availability

Considering the high public interest in research on COVID-19, qualitative data of participants who have indicated their agreement to this as part of the informed consent procedure can be shared with other researchers. However, to preserve the anonymity of respondents and considering the personal nature of qualitative data, requests will be considered on a case-by-case basis. Please contact the corresponding author.

## Acknowledgments

The authors are grateful to Marie A. E. Mueller for her advice on statistical aspects of the project.

## Author contribution

**Alister C. Baird:** Original manuscript formation, Statistical analysis, writing.

**Angela Hassiotis:** Conceptualisation, Review and editing, Supervision.

**Efstathios Papachristou:** Statistical analysis, Supervision, Review and editing.

**Eirini Flouri:** Conceptualization, Methodology, Supervision, Review and editing.

**All authors:** Agree to be accountable for all aspects of the work in ensuring that questions related to the accuracy or integrity of any part of the work are appropriately investigated and resolved.

