## Supplementary Tables S1-S5 for "The Role of Physical Environmental Characteristics and Intellectual Disability in Conduct Problem Trajectories Across Childhood: A Population-Based Cohort Study"

**S1: MCS Multiple imputation results: iteration 1**

***Supplementary Table 1: Greenspace interaction model***

| Table 1: Fully adjusted greenspace model predicting SDQ conduct scores (N = 8168). | | |
| --- | --- | --- |
| Fixed effects | Coefficient (SE) | 95% CI |
| Age | -0.238 (0.008)** | [-0.253, -0.223] |
| Age^2^ | 0.052 (0.001)** | [0.049, 0.054] |
| ID diagnosis | 0.780 (0.327) | [0.140, 1.421] |
| Greenspace (ward decile) | -0.002 (0.009) | [-0.020, 0.015] |
| ID diagnosis x Greenspace x Age | 0.007 (0.010) | [-0.013, 0.027] |
| Greenspace x Age | 0.003 (0.001) | [0.000, 0.005] |
| ID diagnosis x Age | 0.004 (0.050) | [ -0.094, 0.102] |
| ID diagnosis x Greenspace | 0.034 (0.059) | [-0.081, 0.150] |
| Air pollution (NO^2^) | -0.014 (0.009) | [-0.031, 0.004] |
| Damp | 0.144 (0.031)** | [0.084, 0.205] |
| Urban | 0.073 (0.056) | [-0.037, 0.183] |
| Access to garden | -0.162 (0.131) | [-0.418, 0.094] |
| Home spatial density | 0.401 (0.063)** | [ 0.279, 0.525] |
| Maternal psychological distress | 0.650 (0.098)** | [0.457, 0.842] |
| Below poverty line | 0.190 (0.037)** | [ 0.118, 0.263] |
| Atypical family structure | 0.346 (0.040)** | [0.275, 0.416] |
| Male | 0.263 (0.035)** | [0.195, 0.331] |
| ASD | 1.193 (0.312)** | [0.581, 1.804] |
| White ethnicity | 0.169 (0.069) | [0.033, 0.302] |
| Constant | 0.614 (0.180)* | [0.261, 0.970] |
| Random effects | Estimate (SE) | 95% CI |
| Level 3 (ward-level) |  |  |
| Intercept variance | 0.177 (0.022) | [0.139, 0.224] |
| Level 2 (Child level) |  |  |
| Slope (age) variance | 0.139 (0.005) | [0.130, 0.148] |
| Intercept variance | 1.058 (0.018) | [1.024, 1.094] |
| Intercept-slope variance | -0.355 (0.034) | [-0.420, -0.287] |
| *Note.* Age was measured in years and grand mean centered (6.71 yrs).  For fixed effects: **p*<.01, ***p*<.001 | | |

***Supplementary Table 2: NO_2_ interaction model***

| Table 1: Fully adjusted greenspace model predicting SDQ conduct scores (N = 8168). | | |
| --- | --- | --- |
| Fixed effects | Coefficient (SE) | 95% CI |
| Age | -0.219 (0.007)** | [-0.234, -0.205] |
| Age^2^ | 0.052 (0.001)** | [0.049, 0.054] |
| ID diagnosis | 0.892 (0.303)* | [0.299, 1.485] |
| Greenspace (ward decile) | -0.000 (0.009) | [-0.018, 0.017] |
| Air pollution (NO^2^) | -0.013 (0.009) | [-0.031, 0.005] |
| ID diagnosis x Air pollution (NO^2^) x Age | 0.009 (0.009) | [-0.008, 0.026] |
| Air pollution (NO^2^) x Age | -0.001 (0.001) | [-0.003, 0.001] |
| ID diagnosis x Age | -0.027 (0.058) | [-0.142, 0.087] |
| ID diagnosis x Air pollution (NO^2^) | 0.006 (0.044) | [-0.081, 0.093] |
| Damp | 0.144 (0.031)** | [0.084, 0.204] |
| Urban | 0.072 (0.056) | [-0.038, 0.182] |
| Access to garden | -0.164 (0.130) | [-0.419, 0.091] |
| Home spatial density | 0.400 (0.063)** | [0.276, 0.522] |
| Maternal psychological distress | 0.651 (0.098)** | [0.459, 0.848] |
| Below poverty line | 0.190 (0.037)** | [0.118, 0.263] |
| Atypical family structure | 0.346 (0.036)** | [0.275, 0.417] |
| Male | 0.263 (0.035)** | [0.195, 0.331] |
| ASD | 1.175 (0.313)** | [0.561, 1.789] |
| White ethnicity | 0.170 (0.068) | [0.037, 0.304] |
| Constant | 0.602 (0.181) | [0.247, 0.967] |
| Random effects | Estimate (SE) | 95% CI |
| Level 3 (ward-level) |  |  |
| Intercept variance | 0.176 (0.022) | [0.138, 224] |
| Level 2 (Child level) |  |  |
| Slope (age) variance | 0.139 (0.005) | [0.131, 0.148] |
| Intercept variance | 1.058 (0.018) | [1.024, 1.094] |
| Intercept-slope variance | -0.356 (0.034) | [-0.420, -0.288] |
| *Note.* Age was measured in years and grand mean centered (6.71 yrs).  For fixed effects: **p*<.01, ***p*<.001 | | |

***Supplementary Table 3: Garden access interaction model***

| Table 1: Fully adjusted greenspace model predicting SDQ conduct scores (N = 8168). | | |
| --- | --- | --- |
| Fixed effects | Coefficient (SE) | 95% CI |
| Age | -0.241 (0.017)** | [-0.274, -0.207] |
| Age^2^ | 0.052 (0.001)** | [0.049, 0.054] |
| ID diagnosis | 0.825 (0.496) | [-0.148, 1.798] |
| Greenspace (ward decile) | -0.000 (0.009) | [-0.148, 1.798] |
| Air pollution (NO^2^) | -0.014 (0.009) | [-0.031, 0.004] |
| ID diagnosis x Access to garden x Age | -0.085 (0.117) | [-0.315, 0.145] |
| Access to garden x Age | 0.016 (0.017) | [-0.016, 0.049] |
| ID diagnosis x Age | 0.115 (0.114) | [-0.108, 0.339] |
| ID diagnosis x Access to garden | 0.113 (0.493) | [-0.853, 1.079] |
| Access to garden | -0.168 (0.134) | [-0.430, 0.094] |
| Damp | 0.144 (0.031) | [0.084, 0.205] |
| Urban | 0.072 (0.056) | [-0.038, 0.182] |
| Home spatial density | 0.400 (0.063) | [0.276, 0.523] |
| Maternal psychological distress | 0.651 (0.068)** | [0.458, 0.844] |
| Below poverty line | 0.189 (0.037)** | [0.116, 0.262] |
| Atypical family structure | 0.346 (0.036)** | [0.276, 0.417] |
| Male | 0.263 (0.035)** | [0.195, 0.331] |
| ASD | 1.183 (0.311)** | [0.574, 1.793] |
| White ethnicity | 0.170 (0.068) | [0.036, 0.304] |
| Constant | 0.611 (0.182)* | [0.255, 0.968] |
| Random effects | Estimate (SE) | 95% CI |
| Level 3 (ward-level) |  |  |
| Intercept variance | 0.176 (0.022) | [0.138, 0.224] |
| Level 2 (Child level) |  |  |
| Slope (age) variance | 0.139 (0.005) | [0.131, 0.148] |
| Intercept variance | 1.058 (0.018) | [1.024, 1.094] |
| Intercept-slope variance | -0.355 (0.034) | [-0.420, -0.287] |
| *Note.* Age was measured in years and grand mean centered (6.71 yrs).  For fixed effects: **p*<.01, ***p*<.001 | | |

***Supplementary Table 4: Urbanicity and Rurality interaction model***

| Table 1: Fully adjusted greenspace model predicting SDQ conduct scores (N = 8168). | | |
| --- | --- | --- |
| Fixed effects | Coefficient (SE) | 95% CI |
| Age | -0.218 (0.007)** | [-0.231, -0.205] |
| Age^2^ | 0.052 (0.001)** | [0.049, 0.054] |
| ID diagnosis | 1.230 (0.361)* | [0.523, 1.938] |
| Greenspace (ward decile) | -0.000 (0.009) | [-0.018, 0.017] |
| Air pollution (NO^2^) | -0.013 (0.009) | [-0.031, 0.004] |
| ID diagnosis x Urban x Age | 0.032 (0.074) | [-0.113, 0.178] |
| Urban x Age | -0.009 (0.008) | [-0.024, 0.006] |
| ID diagnosis x Age | 0.007 (0.066) | [-0.123, 0.136] |
| ID diagnosis x Urban | -0.368 (0.386) | [-1.125, 0.390] |
| Urban | 0.083 (0.057) | [-0.028, 0.194] |
| Damp | 0.144 (0.031)** | [0.084, 0.204] |
| Access to garden | -0.163 (0.131) | [-0.419, 0.093] |
| Home spatial density | 0.400 (0.063)** | [0.277, 0.523] |
| Maternal psychological distress | 0.651 (0.098)** | [0.458, 0.843] |
| Below poverty line | 0.190 (0.037)** | [0.117, 0.263] |
| Atypical family structure | 0.345 (0.036)** | [0.275, 0.416] |
| Male | 0.263 (0.035)** | [0.195, 0.331] |
| ASD | 1.205 (0.313)** | [0.592, 1.818] |
| White ethnicity | 0.169 (0.069) | [0.035, 0.303] |
| Constant | 0.596 (0.183)* | [0.238, 0.953] |
| Random effects | Estimate (SE) | 95% CI |
| Level 3 (ward-level) |  |  |
| Intercept variance | 0.177 (0.022) | [0.139, 0.224] |
| Level 2 (Child level) |  |  |
| Slope (age) variance | 0.139 (0.005) | [0.130, 0.148] |
| Intercept variance | 1.058 (0.018) | [1.024, 1.094] |
| Intercept-slope variance | -0.356 (0.034) | [-0.420, -0.288] |
| *Note.* Age was measured in years and grand mean centered (6.71 yrs).  For fixed effects: **p*<.01, ***p*<.001 | | |

***Supplementary Table 5: Damp interaction model***

| Table 5: Fully adjusted damp interaction model predicting SDQ conduct scores (N = 8168). | | |
| --- | --- | --- |
| Fixed effects | Coefficient (SE) | 95% CI |
| Age | -0.225 (0.005)** | [-0.235, -0.216] |
| Age^2^ | 0.052 (0.001)** | [0.049, 0.054] |
| ID diagnosis | 1.013 (0.177)** | [0.666, 1.361] |
| Greenspace (ward decile) | -0.000 (0.009) | [-0.018, 0.017] |
| Air pollution (NO^2^) | -0.014 (0.009) | [-0.031, 0.004] |
| ID diagnosis x Damp x Age | -0.060 (0.077) | [-0.211, 0.091] |
| Damp x Age | 0.002 (0.011) | [-0.019, 0.022] |
| ID diagnosis x Age | 0.040 (0.029) | [-0.016, 0.096] |
| ID diagnosis x Damp | -0.531 (0.246) | [-1.014, -0.048] |
| Damp | 0.154 (0.033)** | [0.090, 0.218] |
| Urban | 0.071 (0.056) | [-0.039, 0.180] |
| Access to garden | -0.166 (0.130) | [-0.421, 0.089] |
| Home spatial density | 0.399 (0.063)** | [0.278, 0.522] |
| Maternal psychological distress | 0.650 (0.098)** | [0.458, 0.843] |
| Below poverty line | 0.191 (0.037)** | [0.163, 0.303] |
| Atypical family structure | 0.345 (0.036)** | [0.274, 0.416] |
| Male | 0.263 (0.035)** | [0.195, 0.331] |
| ASD | 1.196 (0.313)** | [0.583, 1.810] |
| White ethnicity | 0.171 (0.068) | [0.037, 0.304] |
| Constant | 0.607 (0,180)* | [0.254, 0.960] |
| Random effects | Estimate (SE) | 95% CI |
| Level 3 (ward-level) |  |  |
| Intercept variance | 0.176 (0.022) | [0.138, 0.224] |
| Level 2 (Child level) |  |  |
| Slope (age) variance | 0.139 (0.005) | [0.130, 0.148] |
| Intercept variance | 1.058 (0.018) | [1.023, 1.093] |
| Intercept-slope variance | -0.357 (0.034) | [-0.421, -0.289] |
| *Note.* Age was measured in years and grand mean centered (6.71 yrs).  For fixed effects: **p*<.01, ***p*<.001 | | |

***Supplementary Table 6: Sensitivity analysis: ethnicity x spatial density x Intellectual disability***

| Table 6: ethnicity x spatial density x intellectual disability sensitivity analysis (N = 8168). | | |
| --- | --- | --- |
| Fixed effects | Coefficient (SE) | 95% CI |
| Age | -0.225 (0.005)** | [-0.234, -0.215] |
| Age^2^ | 0.052 (0.001) ** | [0.049, 0.054] |
| ID diagnosis | 2.656 (0.754)** | [1.178, 4.134] |
| Greenspace (ward decile) | 0.000 (0.008) | [-0.017, 0.017] |
| Air pollution (NO^2^) | -0.013 (0.009) | [-0.030, 0.005] |
| ID diagnosis x ethnicity x spatial density | 0.906 (0.670) | [-0.407, 2.219] |
| Ethnicity x spatial density | 0.419 (0.113)** | [0.197, 0.641] |
| ID diagnosis x ethnicity | -1.061 (0.841) | [-2.710, 0.587] |
| ID diagnosis x spatial density | -1.623 (0.580)* | [-2.760, -0.486] |
| Damp | 0.143 (0.031)** | [0.083, 0.203] |
| Urban | 0.067 (0.056) | [-0.042, 0.176] |
| Access to garden | -0.166 (0.130) | [-0.420, 0.089] |
| Spatial density | 0.0751 (0.102) | [-0.124, 0.274] |
| Maternal psychological distress | 0.653 (0.098)** | [0.461, 0.845] |
| Below poverty line | 0.190 (0.037)** | [0.118, 0.263] |
| Atypical family structure | 0.343 (0.036)** | [0.272, 0.414] |
| Male | 0.262 (0.035)** | [0.194, 0.330] |
| ASD | 1.173 (0.312)** | [0.561, 1.786] |
| White ethnicity | -0.199 (0.122) | [-0.438, 0.041] |
| Constant | 0.903 (0.202)** | [0.508, 1.299] |
| Random effects | Estimate (SE) | 95% CI |
| Level 3 (ward-level) |  |  |
| Intercept variance | 0.173 (0.022) | [0.136, 0.222] |
| Level 2 (Child level) |  |  |
| Slope (age) variance | 0.139 (0.005) | [0.130, 0.148] |
| Intercept variance | 1.056 (0.018) | [1.022, 1.092] |
| Intercept-slope variance | -0.357 (0.034) | [-0.422, -0.289] |
| *Note.* Age was measured in years and grand mean centered (6.71 yrs).  For fixed effects: **p*<.01, ***p*<.001 | | |
